# Multidrug-resistant organism bloodstream infections in solid organ transplant recipients and impact on mortality: a systematic review

**DOI:** 10.1101/2024.07.18.24310675

**Authors:** Alice Liu, Adelaide Dennis, Zarin Fariha, Rekha Pai Mangalore, Nenad Macesic

**Author notes:** Corresponding Author: Dr. Nenad Macesic, Department of Infectious Diseases, The Alfred Hospital and School of Translational Medicine, Monash University, Level 1 Alfred Lane House, Alfred Hospital, 55 Commercial Rd, Melbourne, VIC 3004, Australia.

## Abstract

**Background:** Bloodstream infections (BSI) cause significant morbidity and mortality in solid organ transplant (SOT) recipients. There are few data regarding the contribution of multidrug-resistant organisms (MDROs) to these infections.

**Objectives:** We evaluated the resistance-percentage of MDRO BSIs in SOT recipients and assessed associated mortality.

**Methods:** A systematic review

**Data sources:** MEDLINE and Embase databases up to January 2024.

**Study eligibility criteria:** Studies of adult SOT recipients that quantify MDRO BSI resistance-percentage and/or associated crude mortality. MDROs studied were carbapenem-resistant Enterobacterales (CRE), *Acinetobacter baumannii* (CRAB) and *Pseudomonas aeruginosa* (CRPA), third-generation-cephalosporin-resistant Enterobacterales (3GCR-E), methicillin-resistant *Staphylococcus aureus* (MRSA) and vancomycin-resistant *Enterococcus faecium* (VRE).

**Participants:** Adult SOT recipients with a microbiologically confirmed BSI.

**Interventions:** Not applicable.

**Risk of bias assessment:** Newcastle Ottawa Scale.

**Methods of data synthesis:** MDRO BSI resistance-percentage and mortality outcomes were reported as median (IQR) and crude mortality (%), respectively.

**Results:** Of 945 studies identified, 52 were included. Most were retrospective (41/52) and/or single centre (37/52), and liver transplantation was the most studied SOT type (22/52). High resistance-percentages of BSIs were noted, ranging from 13.6% CRE for Enterobacterales to 59.2% CRAB for *Acinetobacter baumannii*. Resistance-percentage trends decreased over time, but these changes were not statistically significant. Asia had highest resistance-percentages for MRSA (86.2% [IQR 77.3-94.6%]), 3GCR-E (59.5% [IQR 40.5-66.7%]) and CRE (35.7% [IQR 8.3-63.1%]). North America had highest VRE resistance-percentages (77.7% [IQR 54.6-94.7%]). Crude mortality was 15.4-82.4% and was consistently higher than non-MDRO BSIs.

**Conclusions:** MDRO BSIs resistance-percentages were high for all pathogens studied (IQR 24.6-69.4%) but there was geographical and temporal heterogeneity. MDRO BSIs were associated with high mortality in SOT recipients. Microbiological and clinical data in this vulnerable population were incomplete, highlighting the need for robust international multi-centre studies.

## Introduction

Solid organ transplantation (SOT) provides a life-saving measure for patients with end-stage organ failure. However, SOT recipients are at increased susceptibility to infection, which may result in impaired graft function and adverse outcomes (1). Bloodstream infections (BSI) are a leading cause of morbidity and mortality in this population (2–5), with mortality reaching 50% when associated with septic shock (5, 6). Post-transplant BSIs result in prolonged hospital stays, need for re-admission and substantial financial costs (4). Infections with multidrug-resistant organisms (MDROs) are common among SOT recipients due to extended exposure to broad spectrum antimicrobials, immunosuppressive medications, presence of chronic indwelling devices and frequent healthcare exposure (7, 8). SOT itself has been described as an independent risk factor for certain MDRO infections (8). MDRO infections are associated with greater mortality and increased risk of graft failure than those caused by susceptible pathogens (7, 9, 10), posing a major challenge to individual patient care. Furthermore, antimicrobial treatment options for these organisms are often limited and may come with significant toxicities and cost.

While there are multiple reports of high resistance rates and poor outcomes of BSIs in SOT recipients (7, 11–13), there remains an urgent need to better understand the epidemiology and temporal trends in these infections. In the absence of prior systematic review data, we provide a comprehensive description of MDRO BSI resistance-percentages and associated mortality in SOT recipients, focusing on clinically relevant pathogens that influence empirical and directed antimicrobial decision making. The MDROs studied in this review represent a critical group of pathogens that pose a significant global public health threat and are common causes of life-threatening hospital-acquired infection (14).

## Methods

### Study selection

We performed a systematic review of all published literature describing resistance-percentage and/or mortality of BSIs due to MDROs among adult SOT recipients. Six MDROs were selected from the WHO Bacterial Priority Pathogens (2024 update) due to their relevance for SOT recipients (15): carbapenem-resistant Enterobacterales (CRE), carbapenem-resistant *Acinetobacter baumannii* (CRAB), carbapenem-resistant *Pseudomonas aeruginosa* (CRPA), third generation cephalosporin-resistant Enterobacterales (3GCR-E), vancomycin-resistant *Enterococcus faecium* (VRE) and methicillin-resistant *Staphylococcus aureus* (MRSA).

We included all published studies (retrospective and prospective) of adult (≥18 years) SOT recipients (heart, lung, liver, kidney, pancreas, intestinal and multivisceral) that quantified BSIs due to MDROs and/or associated crude mortality rates. Additional studies were identified from review of reference lists. Only full text studies were included in the analysis. Case reports, expert opinion pieces, narrative reviews, pharmacokinetic/pharmacodynamic studies, and studies of colonisation or surveillance blood cultures were excluded. Studies that lacked quantitative data of the proportion of BSIs due to MDROs were also omitted.

The primary outcome was proportion of BSIs due to MDROs, expressed as median [IQR]. The secondary outcome was crude mortality due to MDRO BSI, expressed as a proportion (%).

### Search strategy

A literature search was conducted in the Embase and MEDLINE databases in accordance with guidelines from the Preferred Reporting Items for Systematic Reviews and Meta Analyses (PRISMA) statement (16). No time or language restriction was applied. The following key words and their related MeSH and Emtree search terms were used in various combinations: bloodstream infection, bacteraemia, sepsis, solid organ transplantation, organ graft, multidrug-resistant organism, antimicrobial resistance, carbapenem resistance, extended-spectrum beta-lactamase, vancomycin resistance, methicillin resistance, Enterobacterales, Acinetobacter, Pseudomonas, Staphylococcus and Enterococcus. Full search strategy results are provided in Supp. Tables 1 and 2. The study was registered with PROSPERO (CRD42023466931).

**Table 1:**
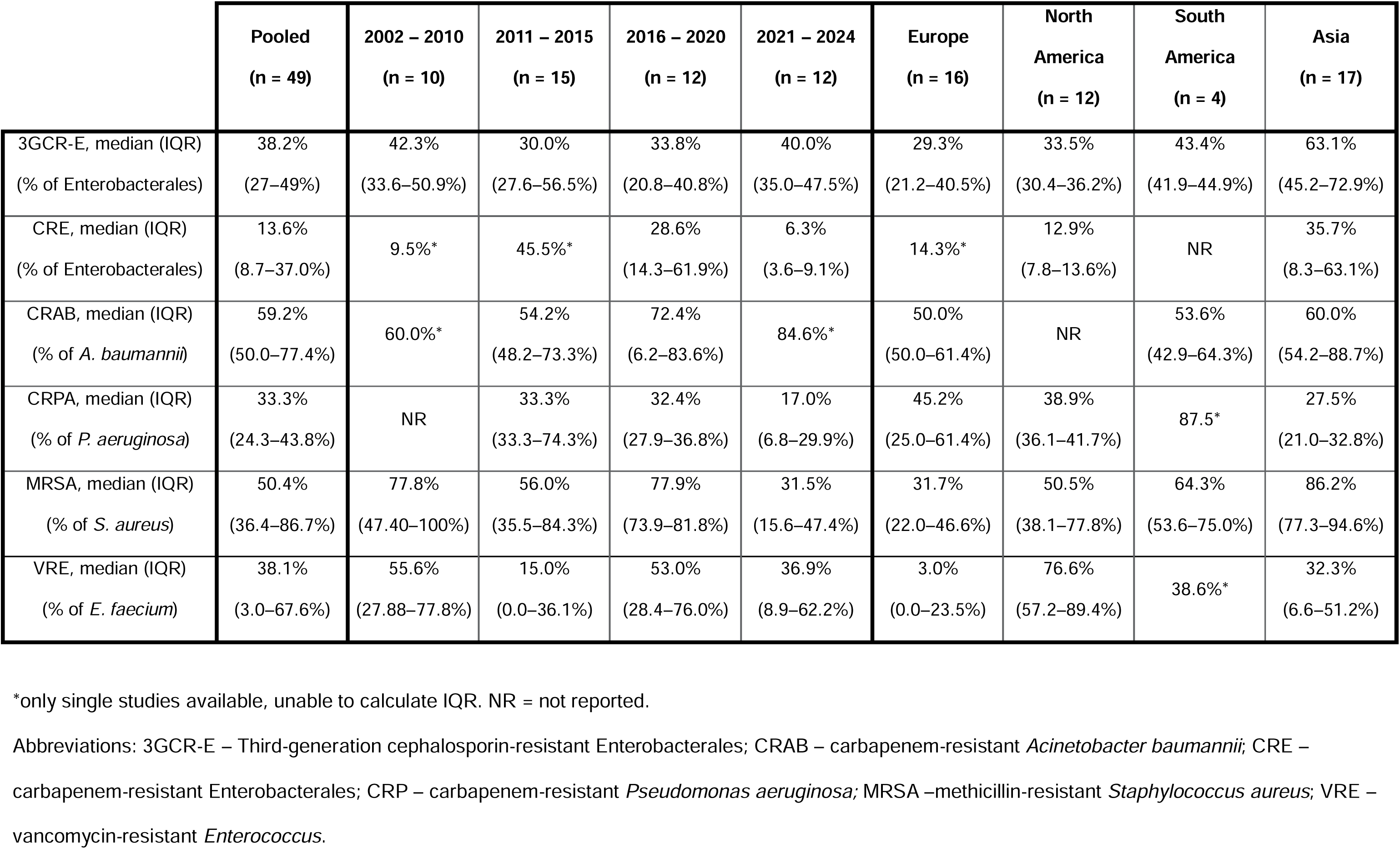
Proportions of bloodstream infections caused by resistant organisms in solid-organ transplant recipients by time period and region.

**Table 2:** Mortality due to bloodstream infection from multidrug resistant organism/s in solid organ transplant recipients:

| Author | Year | Location | Study characteristics | Study population characteristics including SOT type | Organism/s studied | Mortality |
| --- | --- | --- | --- | --- | --- | --- |
| Singh et al. | 2004 | United States | Retrospective, single centre, observational study | 233 liver transplant recipients between 1989 – 2003 | MRSA<br>VRE | MRSA mortality 9/33 (27.3%)<br>VRE mortality 4/8 (50.0%) |
| Husain et al. | 2006 | United States | Prospective, multicentre, observational study (4 sites) | 56 lung transplant recipients with bacteraemia between July 2000 – February 2004 | MRSA | MRSA mortality 1/6 (16.7%) |
| Bedini et al. | 2007 | Italy | Retrospective, single centre, observational study | 205 liver transplant recipients between October 2000 - September 2005 | MRSA | MRSA 30-day mortality 3/5 (60.0%) |
| Kim et al. | 2009 | Korea | Retrospective, single centre, observational study | 144 liver transplant recipients between January 2005 – September 2007 | MRSA<br>VRE<br>ESBL<br>CRAB<br>CRPA | VRE mortality 2/5 (40.0%) |
| Bodro et al. | 2013 | Spain | Prospective, single centre, observational study | 190 SOT recipients (liver, kidney and heart) between January 2007 – December 2012 | MRSA<br>VRE<br>3GCR-E<br>CRAB<br>CRPA | Pooled MDRO mortality 19/54 (35.2%)<br>Pooled non-MDRO mortality 32/185 (14.4%) |
| Aguiar et al. | 2014 | Brazil | Retrospective, single centre, observational study | 759 kidney and 258 liver transplant recipients between January 2000 – September 2008 | 3GCR-E | ESBL 30-day mortality 10/39 (25.6%) |
| Mouloudi et al. | 2014 | Greece | Prospective, single centre, observational study | 17 Liver transplant recipients admitted to the intensive care unit between January 2008 – December 2011 | CRE (CRKP only) | CRKP mortality 14/17 (82.4%) |
| Ye et al. | 2014 | China | Retrospective, multicentre, observational study (2 sites) | 71 SOT recipients between January 2002 – August 2013 | MRSA<br>VRE<br>3GCR-E<br>CRE<br>CRAB | Pooled MDRO 30-day mortality 22/39 (56.4%)<br>Pooled non-MDRO 30-day mortality 12/32 (37.5%) |
| Zhou et al. | 2015 | China | Retrospective, multicentre, observational study (2 sites) | 275 liver transplant recipients with *S. aureus* BSI between January 2001 – December 2014 | MRSA | MRSA mortality 9/20 (45.0%) |
| Kim et al. | 2018 | Korea | Retrospective, single centre, observational study | 393 liver transplant recipients from January 2008 – April 2015 | CRAB | CRAB 1-year mortality 5/14 (35.7%) |
| Kim et al. | 2019 | Korea | Retrospective, single centre, observational study | 536 liver transplant recipients between January 2008 – December 2017 | VRE | VRE 1-year mortality 9/25 (36.0%) |
| Dubler et al. | 2020 | Germany | Retrospective, single centre, observational study | 177 liver transplant recipients between January 2006 – December 2016 who had an *E. faecium* bacteraemia | VRE | VRE 30-day mortality 6/39 (15.4%)<br>VRE 90-day mortality 15/39 (38.5%)<br>Non-VRE 30-day mortality 15/138 (10.9%)<br>Non-VRE 90-day mortality 43/138 (31.2%) |
| Mercuro et al. | 2020 | United States | Retrospective, single centre, cohort study | 58 hospitalised SOT recipients (3 heart, 3 lung, 33 liver, 6 kidney, 10 intestinal, 3 multivisceral) with E faecium BSI between January 2013 – February 2019 | VRE | VRE mortality - 13/41 (31.7%) |
| Kim et al. | 2021 | Korea | Retrospective, single centre, observational study | 727 liver transplant recipients between January 2008 – December 2016 | MRSA<br>VRE<br>3GCR-E<br>CRE<br>CRAB<br>CRPA | Pooled MDRO 90-day mortality 15/39 (38.5%) |
| Lee et al. | 2022 | US | Retrospective, multicentre, cohort study | 117 liver transplant recipients (including 22 multivisceral) between January 2006 – December 2016 | VRE | VRE 1-year mortality 29/108 (26.9%) |
| Anesi et al. | 2023 | United States | Retrospective, multicentre, cohort study | 897 SOT recipients (69 episodes of heart transplant, 58 lung, 278 liver, 514 kidney, 27 pancreas) with an Enterobacterales BSI between 2005 – 2018 | CRE | CRE 60-day mortality 33/70 (47.1%)<br>Non-CRE 60-day mortality 102/827 (12.3%) |
| Min et al. | 2023 | Korea | Retrospective, single centre, nested case control study | 1051 liver transplant recipients between September 2005 – December 2021 | CRAB | CRAB 5-day mortality 17/29 (58.6%)<br>CRAB 10-day mortality 19/29 (65.5%)<br>CRAB 30-day mortality 19/29 (65.5%) |

Abbreviations: 3GCR-E – Third-generation cephalosporin-resistant Enterobacterales, BSI – bloodstream infection, CRAB – carbapenem-resistant *Acinetobacter baumannii*, CRE – carbapenem-resistant Enterobacterales, CRKP – carbapenem-resistant *Klebsiella pneumoniae*, CRPA – carbapenem-resistant *Pseudomonas aeruginosa*, MDRO – multi-drug resistant organism/s, MRSA – methicillin-resistant *Staphylococcus aureus*, SOT – solid organ transplant, VRE – vancomycin-resistant *Enterococcus faecium*.

### Data extraction and analysis

Title and abstract screening were performed by AL. Full text screening was performed by three reviewers (AL, AD, ZF), who also extracted data independently. Conflicts between reviewers on eligibility assessments and data extraction results were resolved through discussion and consensus among all authors. Duplicate studies were removed via Covidence systematic review software (Veritas Health Innovation, Melbourne, Australia) and manual checking. Data were extracted using a custom extraction template covering the following variables: study design, year and location, study population size, type/s of transplant, MDRO species and their frequency, associated crude mortality (where available) and antimicrobial susceptibility testing methods. Statistical analyses and visualisations were performed with R v4.3.0 (2023). Specifically, we applied a linear regression analysis using the ‘lm’ function to assess the presence of a trend in MDRO resistance-percentages over time.

### Risk of bias assessment

Quality assessment was conducted using the Newcastle Ottawa Scale (NOS) (17). For the NOS assessment, exposure was defined as BSI due to MDRO. Comparability of cohorts was based on controlling for baseline patient demographics and immunosuppression regimen. Assessment of MDRO proportions was through microbiological confirmation with a description of the antimicrobial susceptibility methods included. Assessment of mortality outcomes was through an independent assessment or record linkage. There was no minimum follow-up period, and the shortest study period was 12 months (18). Studies with a NOS score of 7 or greater were included in the final analysis.

## Results

### Study characteristics

A total of 1137 studies were identified. After removal of duplicates, 945 were screened, of which 114 studies were retrieved and 52 included in the final analysis (Fig. 1). Reasons for exclusion (62 studies) were incorrect study design, outcomes not specific to BSIs or insufficient data on antimicrobial susceptibility. Most studies were retrospective (41/52, 78.8%) and single-centre (37/52, 71.2%). Of the studies focused on a single transplant type, liver transplant was the most studied (22/52, 42.3%), followed by kidney transplant (5/52, 9.6%). Eighteen (34.6%) studies included pooled multiple transplant types in their analysis. Study population size was variable, ranging from 14 BSI episodes over a 3-year period (19) to 988 episodes over 10 years (8). Full details on the characteristics of the included studies are provided in Supp. Table 3. Quality assessment using the NOS is described in Supp. Tables 4 and 5.

**Fig. 1:**
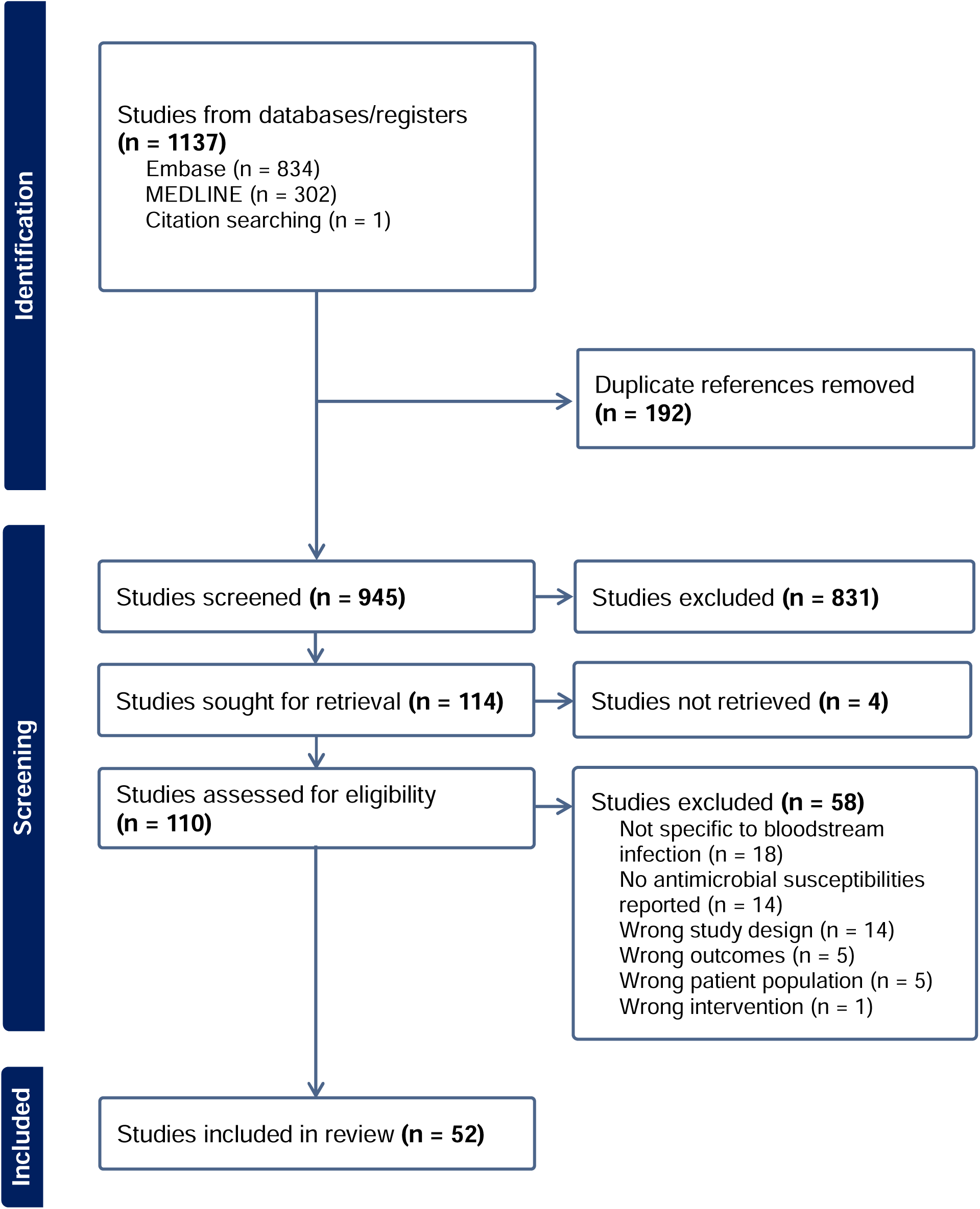
PRISMA flow diagram.

### Pooled estimates of MDRO BSIs in SOT recipients

Proportion of BSIs due to MDROs was reported in 49/52 studies with only crude mortality reported in the remaining three studies. MDRO BSI resistance-percentages are shown in Table 1 and Fig. 2A and ranged from 13.6% (IQR 8.7-37.0%) carbapenem resistance in Enterobacterales to 59.2% (IQR 50.0-77.4%) carbapenem resistance in *Acinetobacter baumannii*. We analysed MDRO resistance-percentages over time and noted reducing trends for all MDROs except CRAB, but these changes were not statistically significant (Table 1, Fig. 2B and Supp. Table 6).

**Fig. 2:**
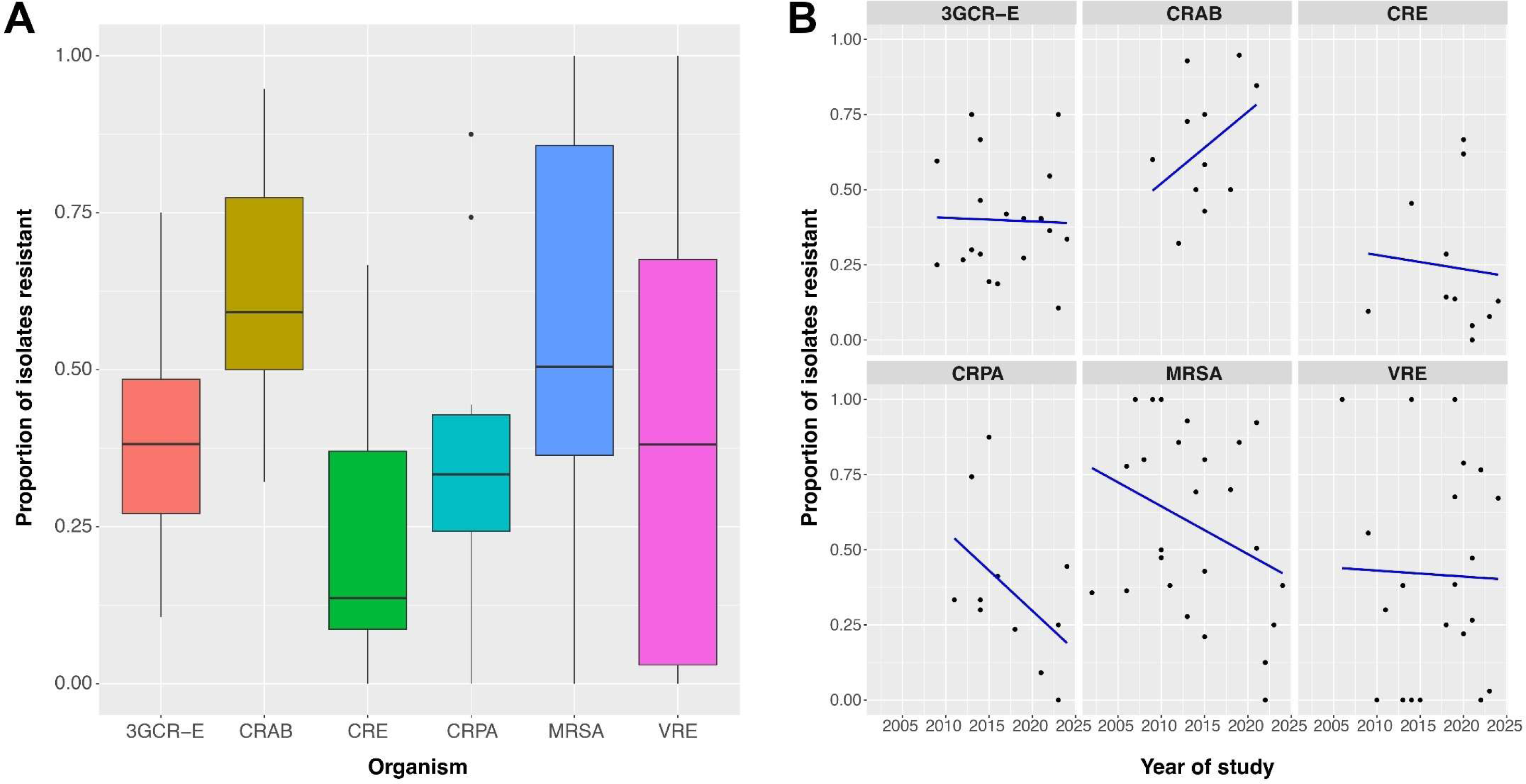
Comparison of proportions of bloodstream infections caused by multidrug-resistant organisms in solid organ transplant recipients by included study. Abbreviations: 3GCR-E – Third-generation cephalosporin-resistant Enterobacterales; CRAB – carbapenem-resistant *Acinetobacter baumannii*; CRE – carbapenem-resistant Enterobacterales; CRP – carbapenem-resistant *Pseudomonas aeruginosa;* MRSA –methicillin-resistant *Staphylococcus aureus*; VRE – vancomycin-resistant *Enterococcus*.

MDRO BSI resistance-percentage varied by region (Table 1), with Asia having highest median resistance-percentages of MRSA (86% [IQR 77-95%]), 3GCR-E (60% [IQR 40-67%]), CRE (36% [IQR 8-63%]) and CRAB (60% [IQR 54-89%]). North America had highest VRE resistance-percentages (median 78% [IQR 55-95%]). South America had highest CRPA resistance-percentage (88%), but this was based on results of a single study.

Liver transplantation was the most studied transplant type with the following median MDRO resistance-percentages: CRE 9.5% (IQR 4.7-14.3%), 3GCR-E 40.4% (IQR 26.7-59.5%), CRAB 72.3% (IQR 48.2-93.3%), CRPA 33.3% (IQR 9.1-5.7%), VRE 34.0% (IQR 24.3-69.8%) and MRSA 80.0% (IQR 48.7-96.4%). Kidney transplantation had sufficient data for analysis of 3GCR-E resistance-percentage (median 32.4% [IQR 21.1-65.3%]), but there were too few studies to determine these for other transplant type/MDRO combinations.

### Pooled estimates of mortality due to MDRO BSI in SOT recipients

Mortality associated with MDRO BSI was reported in 17/52 (32.7%) studies (Table 2). The method of reporting varied between studies, including different time intervals. In most studies (13/17, 76.5%) mortality was recorded by attributable organism, however 3 studies pooled multiple MDRO pathogens for mortality outcomes. Within these limits, crude mortality due to MDRO BSI ranged from 15.4% (VRE 30-day mortality in liver transplant recipients) (20) to 82.4% (CRE mortality in liver transplant recipients admitted to intensive care) (21). Excluding patients in the intensive care setting, crude mortality remained as high as 65.5% (CRAB 10- and 30-day mortality in liver transplant recipients) (22). Where mortality due to MDRO BSI was assessed alongside drug-susceptible BSI, MDRO BSIs were consistently associated with a higher mortality rate irrespective of MDRO or transplant type (Table 2). For individual MDROs, MRSA and VRE-associated mortality were most frequently reported, with a median crude mortality of 36.1% (IQR 19.3–56.3%) and 37.2% (IQR 30.9-42.5%), respectively. There was insufficient data for pooled analysis for all other MDROs studied.

## Discussion

Our systematic review aimed to quantify the burden of MDRO BSIs in SOT recipients and successfully consolidated global antimicrobial resistance (AMR) data for multiple clinically relevant pathogens that cause BSIs in SOT recipients. We observed high resistance-percentages for BSIs in SOT recipients, ranging from median 13.6% for CRE BSIs to median 56.2% for CRAB BSIs. However, there was considerable heterogeneity in resistance-percentages across different regions and time periods. While there were decreasing trends in MDRO BSIs over time, CRAB remained an exception. MDRO BSIs were associated with high crude mortality in SOT recipients, ranging from 15% up to 82.4% in one study in an intensive care setting (21).

We noted prominent differences in MDRO BSI burden across regions. A key question arises regarding comparison with AMR burden in the general population. Asia had highest resistance-percentages for multiple pathogens (MRSA, 3GCR-E, CRE and CRAB). These were higher than in the general hospitalised population in Asia (30.5-49.0% for MRSA BSIs, 36.0-57.2% for 3GCR-E BSIs and 23.1-26.1% for CRE BSIs) (23, 24) but lower for CRAB BSIs (median 60% in SOT recipients vs. 71.5-90.5%) (23, 24). Most Asian studies were confined to liver transplant, which is more commonly associated with post-operative bacterial infections (25–27). This, combined with high overall rates of AMR in many Asian countries may explain the high resistance-percentages for BSIs in SOT recipients we observed in this region (23, 28, 29). North America exhibited the highest resistance percentage of VRE (median 76.6%), aligning with surveillance data demonstrating >70% of US hospital *Enterococcus faecium* isolates are VRE (30). Conversely, Europe had the lowest VRE resistance-percentages (median 3.0%). European general VRE surveillance data range widely from <1% in Holland to 45% in Greece (31). In our review, Spanish (4/16, 25%) and Italian studies (4/16, 25%) were most frequently represented, and our VRE findings reflected surveillance data rates from both countries (3% and 11%, respectively) (31).

Resistance-percentages decreased over time for several of the pathogens studied (MRSA, VRE, 3GCR-E, CRE and CRPA). While these trends did not reach statistical significance, this finding contrasts to global observations of increasing MDRO prevalence over time (32–35). MRSA is unique in demonstrating an early rise and subsequent fall in its prevalence across multiple geographic regions over the last two decades (36), potentially explaining our findings of reducing MRSA trends. The decreasing prevalence of other MDROs over time may potentially be attributed to improvements in infection control measures, better detection of donor-derived infections and changes in antibiotic prophylaxis prescribing practices over time (37, 38). Furthermore, global AMR surveillance methods are not standardised nor is coverage universal (39), and the resistance-percentage rates noted in this study represent specialised transplant centres rather than all healthcare facilities. In contrast, we noted increasing CRAB resistance-percentages in our study, which may reflect the contribution of frequent intensive care unit admission following SOT, with healthcare-associated horizontal transmission of CRAB well described in this setting (40, 41).

We found high crude mortality rates among patients with MDRO BSIs ranging from 15.4 to 82.4%. However, this was based on limited data (17 studies), with almost all studies reporting on <100 SOT recipients with MDRO BSI. These findings contrast with prior reports that noted lower mortality in SOT recipients with sepsis in the early post-transplant period than in non-SOT patients (42, 43). However, these studies did not distinguish between BSI with susceptible and drug-resistant pathogens. Even fewer studies (n=4) compared mortality between MDRO and non-MDRO BSIs in SOT recipients, but in these studies MDRO BSIs were consistently associated with a higher crude mortality rate (10, 20, 44, 45). This may reflect use of empiric antibiotics with inadequate spectra of activity. Alterations in the pathophysiology of different SOT recipients may also account for the high overall mortality rate in this population. For example, heart transplant recipients have higher risk of cardiogenic shock complicating septic shock, while lung transplant recipients are at greater risk of respiratory failure requiring intubation and intensive care support (46).

Our study has several limitations. Firstly, the included studies were heterogeneous in their population size and follow-up period. It was therefore not feasible to conduct a meta-analysis of the study outcomes. The transplant type strongly influences the primary site of infection and associated sepsis (47). We were unable to stratify MDRO frequency-percentages by transplant type because many studies pooled multiple transplant types in their analyses. Additionally, data on prescribing practices were inconsistently recorded across studies and there is a high likelihood that there were geographical and temporal differences in therapeutic and prophylactic antimicrobial prescribing and immunosuppression regimens. These differences would influence MDRO BSI frequency-percentages observed. Finally, mortality data were not reported in a standardised manner, varying from 5-day to 1-year mortality figures, limiting pooled analysis and comparison between studies.

Our review underscores several priorities in the care of SOT recipients. Our findings of high resistance-percentages for BSIs and significant associated mortality highlight that prevention of MDRO acquisition remains paramount. This includes but is not limited to proactive antimicrobial stewardship pre- and post-transplantation to reduce selection pressure, infection control measures to limit cross-transmission in hospital settings, and regular review of immunosuppressant medications and indwelling devices to reduce the risk of colonisation. Strategies for early identification of BSIs caused by MDROs may also help in timely antibiotic rationalisation and limiting the emergence of MDROs in the transplant population, such as continued development of rapid laboratory diagnostics and antimicrobial susceptibility testing methods, and implementation of protocolised surveillance testing programs for donors and recipients.

We also identified future research priorities. Our review found a paucity of transplant type-specific microbiological data, non-standardised mortality reporting methods and a reliance on retrospective and/or single centre data resulting in wide variation in study size. It has been noted that frequency measures such as resistance-percentages are of limited use without accompanying population resistance-percentage estimates or incidence-density data (48), which was also evident in our review. Future work should focus on longitudinal prospective data collection using consistent metrics, capturing both clinical and microbiological outcomes and representing SOT types proportionately. This presents an opportunity to further develop and implement larger scale transplant registries such as the Swiss Transplant Cohort study (11, 13) which capture infectious complications and corresponding microbiological data.

## Conclusion

Our findings reinforce that SOT recipients are highly vulnerable to MDRO BSIs and experience substantial associated mortality. We identified marked geographical variation in the proportion of MDRO BSIs, which warrants ongoing characterisation in future studies. While there was a trend towards reducing resistance-percentages of several MDROs, this was not statistically significant and certain pathogens were under-represented. We noted gaps in the reporting of transplant type-specific data and a paucity of standardised mortality outcomes. Our study highlights the urgent need for robust international multi-centre cohort studies that adequately capture microbiological data and BSI clinical outcomes to guide prevention and empirical treatment strategies in this highly vulnerable patient group.

## Data Availability

All data produced in the present study are available upon reasonable request to the authors

## Transparency declarations

None to declare

