## Supplementary material for "Multidrug-resistant organism bloodstream infections in solid organ transplant recipients and impact on mortality: a systematic review": Supp. Table

**Supplementary Appendix**

**Supp. Table 1 – Embase search strategy results**

Embase Classic+Embase <1947 to 2024 February 19>

| **#** | **Query** | **Results from 20 Feb 2024** |
| --- | --- | --- |
| 1 | transplantation/ or allograft/ or allotransplantation/ | 282,774 |
| 2 | graft recipient/ or engraftment/ | 100,816 |
| 3 | organ transplantation/ or tissue transplantation/ or tissue graft/ or heart transplantation/ or heart graft/ or heart lung transplantation/ or intestine transplantation/ or intestine graft/ or kidney transplantation/ or cadaver kidney/ or kidney allograft/ or kidney autotransplantation/ or kidney graft/ or kidney pancreas transplantation/ or liver transplantation/ or lung transplantation/ or hepatocyte transplantation/ or kidney pancreas transplantation/ or orthotopic transplantation/ or pancreas transplantation/ or pancreas islet transplantation/ or heterotopic transplantation/ or spleen transplantation/ | 728,631 |
| 4 | patient history of transplantation/ or patient history of heart transplantation/ or patient history of kidney transplantation/ or patient history of liver transplantation/ or patient history of lung transplantation/ | 3,989 |
| 5 | living donor/ | 56,524 |
| 6 | ((SOT or organ* or transplant* or graft*) adj recipient*).mp. | 200,103 |
| 7 | ((organ* or visceral or multivisceral or heart* or cardiac* or cardiopulmonary* or intestin* or bowel* or kidney* or renal* or nephr* or liver* or hepat* or lung* or pulmonary or pancrea* or pancreas kidney or orthotopic or heterotopic) adj3 (transplant* or graft* or allograft*)).mp. | 922,529 |
| 8 | ((cardiac* or heart* or renal* or kidney* or nephr* or lung* or pulmonary or liver* or hepatic* or pancrea* or intestin* or small bowel* or visceral or multivisceral) adj recipient*).mp. | 15,618 |
| 9 | 1 or 2 or 3 or 4 or 5 or 6 or 7 or 8 | 1,167,488 |
| 10 | bloodstream infection/ or bacteremia/ or sepsis/ or septic shock/ or septicemia/ or toxic shock syndrome/ or pyemia/ | 465,036 |
| 11 | Gram negative sepsis/ or staphylococcal bacteremia/ | 7,472 |
| 12 | (septic or septic?emi* or septicopy?emi* or pyoseptic?emi* or pyoh?emi* or py?emi* or BSI or bacter?emi* or bacill?emi*).mp. | 394,777 |
| 13 | (tox?emi* or endotox?emi* or blood poisoning or bloodstream poison*).mp. | 50,530 |
| 14 | (infection* adj3 (blood or bloodstream)).mp. | 73,628 |
| 15 | ((toxin* or endotoxin*) adj3 (blood or bloodstream)).mp. | 3,514 |
| 16 | 10 or 11 or 12 or 13 or 14 or 15 | 686,715 |
| 17 | drug resistance/ or multidrug resistance/ or antibiotic resistance/ | 459,455 |
| 18 | carbapenem resistance/ or carbapenem resistant Acinetobacter baumannii/ or carbapenem-resistant Enterobacteriaceae/ or carbapenem resistant Escherichia coli/ or carbapenem resistant Klebsiella pneumoniae/ or carbapenem resistant Pseudomonas aeruginosa/ or carbapenemase/ or carbapenemase producing Enterobacteriaceae/ or multidrug resistant Escherichia coli/ or extended spectrum beta lactamase/ or extended spectrum beta lactamase producing Enterobacteriaceae/ or extended spectrum beta lactamase producing Escherichia coli/ or extended spectrum beta lactamase producing Klebsiella pneumoniae/ or vancomycin resistance/ or vancomycin intermediate Staphylococcus aureus/ or vancomycin resistant Staphylococcus aureus/ or methicillin resistance/ or methicillin resistant Staphylococcus aureus/ or methicillin resistant Staphylococcus aureus bacteremia/ or methicillin resistant Staphylococcus aureus infection/ or vancomycin resistant Enterococcus/ or vancomycin-resistant Enterococcus infection/ or Acinetobacter/ or Acinetobacter baumannii/ or Acinetobacter infection/ or extensively drug resistant Acinetobacter baumannii/ or multidrug resistant Acinetobacter baumannii/ or Acinetobacter baumannii-calcoaceticus complex infection/ or Citrobacter/ or Citrobacter freundii/ or Citrobacter rodentium/ or Citrobacter amalonaticus/ or Citrobacter braakii/ or Citrobacter koseri/ or Citrobacter intermedius/ or Enterobacter/ or Enterobacter aerogenes/ or Enterobacter cloacae/ or Enterobacter asburiae/ or Escherichia coli/ or Morganella/ or Morganella morganii/ or Proteus mirabilis/ or Proteus vulgaris/ or Proteus mirabilis infection/ or Proteus penneri/ or Proteus infection/ or Providencia/ or Serratia/ or Serratia plymuthica/ or Serratia infection/ or Serratia liquefaciens/ | 932,800 |
| 19 | (MDR or CRE or Carbepenemase-producing or ESBL or MRSA or VISA or VRSA or VRE).mp. | 256,573 |
| 20 | ((Antibiotic* or antibacterial* or antimicrobial* or multiple drug* or multidrug) adj3 (resistan* or non-susceptible or nonsusceptible)).mp. | 553,699 |
| 21 | ((carbapenem or beta lactam* or vancomycin or VAN or methicillin) adj (resistan* or non-susceptible or nonsusceptible)).mp. | 169,997 |
| 22 | (resistant adj (Acinetobacter or Pseudomonas or Citrobacter or Enterobacter or Escherichia or Klebsiella or Morganella or Proteus or Providencia or Serratia or Staphylococcus or Enterococc*)).mp. | 154,337 |
| 23 | 17 or 18 or 19 or 20 or 21 or 22 | 1,655,826 |
| 24 | 9 and 16 and 23 | 3,325 |
| 25 | (exp animal/ or exp invertebrate/ or animal.hw. or nonhuman/) not exp human/ | 13,389,423 |
| 26 | 24 not 25 | 3,265 |
| 27 | limit 26 to (embryo or infant or child or preschool child <1 to 6 years> or school child <7 to 12 years> or adolescent <13 to 17 years>) | 965 |
| 28 | 26 not 27 | 2,300 |
| 29 | (child* or adoles* or teen* or p?ediatric* or toddler* or infan* or newborn* or neonat* or preschool* or pre-school* or schoolchild* or primary school* or secondary school* or kinder*).mp. | 10,510,835 |
| 30 | 28 not 29 | 2,140 |
| 31 | limit 30 to (books or chapter or conference abstract or erratum or letter or note or short survey or tombstone) | 938 |
| 32 | 30 not 31 | 1,202 |
| 33 | (case report* or editorial* or modelling).mp. | 7,465,120 |
| 34 | 32 not 23 | 834 |

**Supp. Table 2 – MEDLINE search strategy results**

Ovid MEDLINE(R) ALL <1946 to February 19, 2024>

| **#** | **Query** | **Results from 20 Feb 2024** |
| --- | --- | --- |
| 1 | transplants/ or allografts/ | 248,303 |
| 2 | Transplant Recipients/ | 82,707 |
| 3 | transplantation/ or cell transplantation/ or organ transplantation/ or heart transplantation/ or kidney transplantation/ or liver transplantation/ or lung transplantation/ or pancreas transplantation/ or transplantation, heterotopic/ | 824,895 |
| 4 | Living Donors/ | 50,827 |
| 5 | ((SOT or organ* or transplant* or graft*) adj recipient*).mp. | 200,103 |
| 6 | (transplant* medicine or living donor* or allotransplant* or engraftment).mp. | 184,164 |
| 7 | ((organ* or cardiac* or heart* or renal* or kidney* or nephr* or lung* or pulmonary or liver* or hepatic* or pancrea* or intestin* or small bowel* or visceral or multivisceral) adj3 (transplant* or graft* or allograft*)).mp. | 907,487 |
| 8 | ((cardiac* or heart* or renal* or kidney* or nephr* or lung* or pulmonary or liver* or hepatic* or pancrea* or intestin* or small bowel* or visceral or multivisceral) adj recipient*).mp. or multidrug-resistant organisms/ | 15,619 |
| 9 | 1 or 2 or 3 or 4 or 5 or 6 or 7 or 8 | 1,205,094 |
| 10 | Bacteremia/ or Sepsis/ or Hemorrhagic Septicemia/ or Shock, Septic/ or Toxemia/ or Endotoxemia/ | 431,318 |
| 11 | (septic* or BSI or bacter?emi*).mp. | 397,194 |
| 12 | (Tox?emi* or Endotox?emi* or blood* poisoning or blood stream poisoning).mp. | 50,530 |
| 13 | (infection* adj3 (blood or bloodstream)).mp. | 73,628 |
| 14 | ((toxin* or endotoxin*) adj3 (blood or bloodstream)).mp. | 3,514 |
| 15 | 10 or 11 or 12 or 13 or 14 | 684,313 |
| 16 | Microbial Sensitivity Tests/ or Drug Resistance, Multiple/ or Drug Resistance, Multiple, Bacterial/ or Drug Resistance, Bacterial/ or Drug Resistance, Microbial/ or multidrug-resistant organisms/ | 517,032 |
| 17 | Carbapenem-Resistant Enterobacteriaceae/ or beta-Lactam Resistance/ or Acinetobacter baumannii/ or Acinetobacter Infections/ or Pseudomonas Infections/ or Pseudomonas aeruginosa/ or Enterobacteriaceae/ or Enterobacteriaceae Infections/ or Citrobacter/ or Citrobacter freundii/ or Citrobacter koseri/ or Enterobacter aerogenes/ or Enterobacter cloacae/ or Escherichia coli/ or Escherichia coli Infections/ or Klebsiella oxytoca/ or Klebsiella pneumoniae/ or Klebsiella Infections/ or Morganella morganii/ or Proteus mirabilis/ or Proteus penneri/ or Proteus vulgaris/ or Proteus Infections/ or Serratia/ or Serratia marcescens/ or Serratia Infections/ or Methicillin Resistance/ or Methicillin-Resistant Staphylococcus aureus/ or Vancomycin-Resistant Staphylococcus aureus/ or Staphylococcal Infections/ or Vancomycin Resistance/ or Enterococcus/ or Enterococcus faecium/ or Vancomycin-Resistant Enterococci/ | 1,192,054 |
| 18 | (multidrug resistan* or antimicrobial resistan* or anti-microbial resistan* or multipl* resistan* or MDR or drug resistan* or carbapenem-resistan* or carbapenemase or extended-spectrum beta-lactamase or ESBL or MRSA or methicillin-resistant or VRSA or VISA or vancomycin-intermediate S aureus or vancomycin-intermediate Staph aureus or VRE or vancomycin resistan*).mp. | 1,025,186 |
| 19 | (resistant adj (Acinetobacter or Pseudomonas or Enterobacter or Citrobacter or Escherichia or Klebsiella or Morganella or Proteus or Serratia or Staphylococcus aureus or Enterococc*)).mp. | 151,676 |
| 20 | 16 or 17 or 18 or 19 | 2,128,375 |
| 21 | 9 and 15 and 20 | 4,439 |
| 22 | exp animals/ not humans.sh. | 38,706,384 |
| 23 | 21 not 22 | 1,002 |
| 24 | limit 23 to ("all infant (birth to 23 months)" or "all child (0 to 18 years)" or "newborn infant (birth to 1 month)" or "infant (1 to 23 months)" or "preschool child (2 to 5 years)" or "child (6 to 12 years)" or "adolescent (13 to 18 years)") | 350 |
| 25 | 23 not 24 | 652 |
| 26 | (child* or adoles* or teen* or p?ediatric* or toddler* or infan* or newborn* or neonat* or preschool* or pre-school* or schoolchild* or primary school* or secondary school* or kinder*).mp. | 10,510,835 |
| 27 | 25 not 26 | 634 |
| 28 | (pregnan* or gestation* or antenatal or prenatal or natal or gravidit* or gravida* or multigravid* or primigravid*).mp. | 2,855,753 |
| 29 | 27 not 28 | 631 |
| 30 | limit 29 to (address or autobiography or bibliography or biography or case reports or congress or consensus development conference or consensus development conference, nih or duplicate publication or "expression of concern" or interactive tutorial or interview or lecture or legal case or legislation or letter or newspaper article or patient education handout or periodical index or personal narrative or randomized controlled trial, veterinary or "research support, american recovery and reinvestment act" or research support, nih, extramural or research support, nih, intramural or research support, non us gov't or research support, us gov't, non phs or research support, us gov't, phs or retracted publication or "retraction of publication" or technical report or video-audio media or webcast) | 313 |
| 31 | 29 not 30 | 318 |
| 32 | (address or autobiography or bibliography or biography or case report or clinical conference* or comment* or editorial* or guideline* or interactive tutorial* or interview* or lecture* or legal case* or legislation* or newspaper article* or patient education handout* or personal narrative* or portrait or practice guideline* or technical report* or video-audio media* or webcast* or conference abstract* or letter* or book* or chapter* or modelling).mp. | 14,327,520 |
| 33 | 31 not 32 | 302 |

**Supp. Table 3 – Resistance-percentages for multidrug resistant organisms (MDRO) bloodstream infections (BSI) for all included studies.**

| **Author** | **Study type** | **Location** | **Study population characteristics** | **Organism/s studied** | **MDRO prevalence** | **MDRO Mortality** | **Microbiological laboratory methods** |
| --- | --- | --- | --- | --- | --- | --- | --- |
| Adelman et al. (2024) | Retrospective, single centre, observational study | United States | 2293 SOT recipients (1251 kidney, 663 liver, 219 heart, 160 multivisceral) with BSI within the first year of transplant, between June 2016 - September 2021. | MRSA  VRE  3GCR-E  CRE  CRPA | 313 episodes of BSI among 196 patients  MRSA 8/21 (38.1%)  VRE 45/67 (67.2%)  3GCR-E 52/155 (33.6%)  CRE 20/155 (12.9%)  CRPA 12/27 (44.4%) | Not reported | Microbiological identification was performed using MALDI-TOF and AST determined with BD Phoenix Automated System. Results were interpreted according to CLSI criteria. |
| Aguiar et al.  (2014) | Retrospective, single centre, observational study | Brazil | 759 kidney and 258 liver transplant recipients between January 2000 - September 2008 | 3GCR-E | 112 episodes of BSI due to Enterobacterales  3GCR-E 52/112 (46.2%) | ESBL 30-day mortality 10/39 (25.6%)  ESBL 30-day mortality after recurrent bacteraemia 2/39 (5.1%) | Microbiological identification and AST were performed by using the Vitek-2 system. Results were interpreted according to CLSI criteria. |
| Anesi et al. (2021) | Retrospective, multicentre, case control study | United States | 897 SOT recipients (77 heart, 65 lung, 310 liver, 585 kidney, 27 pancreas) with Enterobacterial BSI between January 2005 - December 2015 | 3GCR-E | 3GCR-E 395/988 (40.0%) | Not reported | Microbiological identification and AST were performed using either Vitek-2 semiautomated system or BD Phoenix Automated System or disc diffusion method (prior to 2010). |
| Anesi et al. (2023) | Retrospective, multicentre, cohort study | United States | 897 SOT recipients (69 episodes of heart transplant, 58 lung, 278 liver, 524 kidney, 27 pancreas) with an Enterobacterales BSI between January 2005 - January 2018 | CRE | CRE 70/897 (7.8%) | CRE 60-day mortality 33/70 (47.1%)  Non-CRE 60-day mortality 102/827 (12.3%) | Microbiological identification and AST were performed using either Vitek-2 semiautomated system or BD Phoenix Automated System or disc diffusion method (prior to 2010). |
| Basaran et al. (2022) | Retrospective, single centre,  cohort study | Turkey | 702 kidney transplant recipients between January 2000 - January 2016 | MRSA  VRE  3GCR-E | Total 74 episodes of BSI among 56 patients  MRSA = 1/8 (12.5%)  VRE = 0/4 (0%)  3GCR-E = 16/44 (36.4) | Not reported | Between 2000 and 2014, microbiological identification and AST were performed using the BD Phoenix automated system. Between January 2014 and November 2017, the Vitek-2 automated system was used. Interpretation methods not stated. |
| Bedini et al. (2007) | Retrospective, single centre, observational study | Italy | 205 Liver transplant recipients between October 2000 - September 2005 | MRSA | 42 episodes of BSI among 28 patients  MRSA 5/5 (100%) | Not reported | Not reported |
| Berger et al. (2006) | Retrospective,  single centre, cohort study | Austria | 217 kidney-pancreas transplant recipients between March 1997 - Oct 2004 | MRSA | 39 patients with BSI  MRSA 4/11 (36.4%) | Not reported | Not reported |
| Bert et al. (2010) | Retrospective, single centre, cohort study | France | 704 liver transplant recipients between January 1997 - December 2007 | MRSA | 259 episodes of BSI among 205 patients  MRSA 28/56 (50%) | Not reported | Not reported |
| Bodro et al. (2013) | Prospective, single centre, observational study | Spain | 190 SOT recipients (liver, kidney and heart) between January 2007 - December 2012 | MRSA  VRE  3GCR-E  CRAB  CRPA | 276 episodes of BSI  MRSA 5/18 (27.8%)  VRE 0/16 (0%)  3GCR-E 15/50 (30.0%)  CRAB 8/11 (72.7%)  CRPA 26/35 (74.3%) | Pooled MDRO mortality 19/54 (35.2%)  Pooled non-MDRO mortality 32/185 (14.4%) | Microbiological confirmation and AST were performed using commercial panels from the MicroScan automated system.  Results were interpreted according to CLSI criteria. |
| Camargo et al. (2015) | Prospective, multicentre. observational study (16 sites) | Brazil | 87 SOT transplant recipients (20 liver, 7 lung, 45 kidney, 2 heart, 2 pancreas, 5 multivisceral, 6 not specified) between June 2007 - 31 March 2010 | MRSA  CRAB  CRPA | Total 83 episodes of monomicrobial BSI  MRSA 6/14 (42.9%)  CRAB 3/4 (75%)  CRPA 7/8 (87.5%) | Not reported | Microbiological identification and AST were performed using standard methods and results were interpreted according to CLSI criteria. |
| Chueiri Neto et al. (2019) | Retrospective, single centre observational study | Brazil | 401 Liver transplant recipients between January 2005 - June 2016 who developed an early post-operative bloodstream infection | MRSA VRE 3GCR-E | 139 episodes of BSI among 103 patients  MRSA 6/7 (85.7%)  VRE 5/6 (83.3%)  3GCR-E 19/47 (40.4%) | Not reported | Microbiological identification and antimicrobial susceptibility testing methods not described |
| Dasakalaki et al. (2014) | Retrospective, single centre, observational study | Greece | 108 kidney transplant recipients between January 2010 - May 2013 | MRSA  VRE  3GCR-E  CRAB  CRPA | 26 episodes of BSI among 22 patients  MRSA 1/1 (100%)  VRE 2/2 (100%)  3GCR-E 4/14 (28.6%)  CRAB 1/2 (50%)  CRPA 1/3 (33.3%) | Not reported | ESBL production was confirmed with the double disc synergy test according to CLSI criteria. Carbapenemase production was confirmed using modified Hodge’s method |
| De Gouvea et al. (2012) | Retrospective single centre, observational study | Brazil | 562 kidney and 238 liver transplant recipients between January 2002 to January 2009 | CRAB | 28 episodes of A baumannii BSI (16 in liver transplant patients, 12 in kidney transplant patients)  CRAB 9/28 (32.1%) | Not reported | Microbiological identification and AST were performed using the Vitek-1 system with additional susceptibilities via disc diffusion according to CLSI methods.  From July 2006 onwards, isolates intermediate to imi-/meropenem defined as carbepenem resistant |
| Dubler et al. (2020) | Retrospective, single centre, observational study | Germany | 177 liver transplant recipients between January 2006 - December 2016 who had an Enterococcus faecium BSI | VRE | Total 177 episodes of E faecium BSI among liver transplant recipients  VRE = 39/177 (22.0%) | VRE 30-day mortality 6/39 (15.4%)  VRE 90-day mortality 15/39 (38.5%)  Non-VRE 30-day mortality 15/138 (10.9%)  Non-VRE 90-day mortality 43/138 (31.2%) | Real time PCR was used for identification of vanA and vanB isolates. Results were interpreted according to EUCAST criteria. |
| Eichenberger et al. (2021) | Prospective, multicentre, cohort study (2 sites) | United States | 103 SOT recipients (18 heart, 39 lung, 9 liver, 26 kidney, 11 multivisceral) with S. aureus BSI between January 2005 - December 2019 | MRSA | MRSA 52/103 (50.5%) | Not reported | AST was performed using the MicroScan Walkaway system  (microbroth dilution method).  Results were interpreted according to CLSI guidelines.  Spa genotyping was performed using RidomStaphType. |
| Gagliotti et al. (2018) | Prospective, multicentre cohort study (10 centres) | Italy | 113 lung and 523 liver recipients between January 2014 - January 2015 | MRSA  VRE  CRE  CRAB  CRPA | Lung transplant recipients - 15 episodes of BSI:  CRE - 2/7 (28.6%)  CRAB 1/1 (100%)  CRPA 1/1 (100%)  Liver transplant recipients - 84 episodes of BSI  MRSA 7/10 (70.0%)  VRE 2/8 (25.0%)  3GCR-E 15/35 (42.9%)  CRE - 5/35 (14.3%)  CRAB 1/2 (50.0%)  CRPA 4/7 (57.1%) | Not reported | Not reported |
| Gao et al. (2015) | Retrospective, multicentre, observational study (2 sites) | China | 17 liver transplant recipients who developed A. baumannii infection between January 2007 - December 2014 | CRAB | CRAB 3/7 (42.9%) | Not reported | Microbiological identification was performed using the Vitek-2 system AST was performed using the Kirby-Bauer method and MIC tests according to CLSI criteria. |
| Hashimoto et al. (2008) | Retrospective, single centre, observational study | Japan | 242 liver transplant recipients with BSI between January 1996 - November 2004. | MRSA | 25 episodes of bacteraemia  MRSA 4/5 (80%) | Not reported | Not reported |
| Husain et al. (2006) | Prospective, multicentre, observational study (4 sites) | United States | 56 lung transplant recipients with bacteraemia between July 2000 - February 2004 | MRSA  VRE | MRSA 7/9 (77.8%)  VRE 3/3 (100%) | MRSA mortality 1/6 (16.7%) | Not reported |
| Iida et al. (2010) | Retrospective, single centre, observational study | Japan | 181 liver transplant recipients between April 2006 – November 2009 | MRSA | MRSA 9/19 (47.7%) | Not reported | Not reported |
| Lee et al. (2011) | Retrospective, single centre, cohort study | United States | 737 liver transplant recipients between January 1997 - March 2006 | MRSA  VRE  CRPA | 123 LTx patients who developed BSI  MRSA 8/21 (38.1%)  VRE 3/10 (30%)  CRPA 3/9 (33.3%) | Not reported | Microbiological identification and AST performed using standard techniques according to CLSI lab methods. |
| Karruli et al. (2021) | Retrospective, single centre observational study | Italy | 47 orthoptic heart transplant recipients between January 2016 - December 2018 | MRSA  VRE | 14 episodes of BSIs  MRSA 1/1 (100%)  VRE 0/1 (0%) | Not reported | Microbiological identification and AST were performed using the Vitek-2 automated system. Results were interpreted according to EUCAST criteria. |
| Kim et al. (2009) | Retrospective, single centre, observational study | Korea | 144 liver transplant recipients between January 2005 - September 2007 | MRSA  VRE  3GCR-E  CRAB  CRPA | 40 episodes of BSI among 34 patients  MRSA 3/3 (100%)  VRE 5/9 (54.5%)  ESBL - 2/8 (25%)  CRAB 0/1 (0%)  CRPA 0/1 (0%) | VRE mortality 2/5 (40%) | ot reported |
| Kim et al. (2013) | Retrospective, single centre, observational study | Korea | 222 liver transplant recipients with BSI between February 2005 - May 2011 | MRSA  VRE  3GCR-E  CRAB | 112 episodes of BSI among 64 patients  MRSA 13/14 (92.8%)  VRE 8/21 (38%)  3GCR-E 27/36 (75%)  CRAB 13/14 (92.8%) | Not reported | AST was performed using the minimal inhibitory concentration agar dilution method according to CLSI criteria. |
| Kim et al. (2018) | Retrospective, single centre, observational study | Korea | 393 liver transplant recipients between January 2008 - April 2015 | CRAB | CRAB 18/19 (94.7%) | CRAB 1-year mortality 5/14 (35.7%) | Microbiological and AST were performed using VITEK-2 automated system. Results were interpreted according to CLSI criteria.  Carbapenem resistance was defined as non-susceptible to mero-/ ertapenem and/or imipenem in vitro. Isolates that were intermediate on testing were considered resistant. |
| Kim et al. (2019) | Retrospective, single centre, observational study | Korea | 536 liver transplant recipients between January 2008 -December 2017 | VRE | 58 episodes of enterococcal BSI among 42 patients:  VRE 25/37 (67.5%) | VRE 1-year mortality 9/25 (36%) | Microbiological identification performed using VITEK 2 automated system. Susceptibility results interpreted according to CLSI criteria. |
| Kim et al. (2021) | Retrospective, single centre, observational study | Korea | 727 liver transplant recipients between January 2008 - December 2016 | MRSA  VRE  3GCR-E  CRE  CRAB  CRPA | 149 episodes of BSI among 108 patients  MRSA 12/13 (92.3%)  VRE 17/64 (26.6%)  3GCR-E 17/42 (40.5%)  CRE 2/42 (4.8%)  CRAB 11/13 (84.6%)  CRPA 1/11 (9.1%) | Pooled MDRO 90-day mortality 15/39 (38.5%) | Microbiological identification and AST was performed using the Vitek-2 automated systemic. Results were interpreted according to CLSI criteria. |
| Lee et al. (2022) | Retrospective, multicentre, cohort study (9 sites) | United States | 117 liver transplant recipients (including 22 multivisceral) between January 2006 – December 2016 | VRE | VRE 108/141 (78%) | VRE 1-year mortality 29/108 (26.9%)  VRE 30-day mortality from first bacteraemia 24/108 (22.2%)  VRE 30-day mortality from last bacteraemia 31/108 (28.7%). | Microbiological identification data were provided by participating clinical laboratories). AST was performed and interpreted interpreted according to CLSI criteria. |
| Luo et al. (2016) | Retrospective, multicentre, observational study (2 sites) | China | 1967 SOT recipients (1646 kidney, 310 liver, 3 heart, 8 multivisceral) between January 2003 - July 2015 | CRPA | CRPA 7/17 (41.2%) | Not reported | Microbiological identification was performed using the Vitek-2 automated system. AST was determined using the Kirby-Bauer method and MIC testing. Results were interpreted according the NCCLs manual in 2003 and CLSI criteria from 2004 - 2015. |
| Malinis et al. (2012) | Retrospective, single centre, case control analysis | United States | 3047 SOT recipients (485 lung, 716 liver, 1299 kidney, 588 heart, 54 pancreas, 4 small intestine) between January 2000 – December 2008  Subset of 70 who were SOT recipients (26 lung, 19 liver, 18 kidney, 7 heart) - case controlled with non-transplant recipients | MRSA | MRSA 60/70 (85.7%) | Not reported | Not reported |
| Mercuro et al. (2020) | Retrospective, cohort study, single centre | United States | 58 hospitalised SOT recipients (3 heart, 3 lung, 33 liver, 6 kidney, 10 intestinal, 3 multivisceral) with E faecium BSI between January 2013 - February 2019 | VRE | VRE 41/52 (78.8%) | VRE mortality - 13/41 (31.7%) | Microbiological identification and AST were performed using Vitek-2. Results were interpreted according to CLSI guidelines. |
| Min et al. (2023) | Retrospective, single centre, nested case control study | Korea | 1051 liver transplant recipients between September 2005 – December 2021 | CRAB | Not reported | CRAB 5-day mortality 17/29 (58.6%)  CRAB 10-day mortality 19/29 (65.5%)  CRAB 30-day mortality 19/29 (65.5%) | Not reported |
| Mouloudi et al. (2014) | Prospective, single centre, observational study | Greece | 17 Liver transplant recipients admitted to the intensive care unit between January 2008 – December 2011 | CRE (K. pneumoniae only) | Not reported | CRKP mortality 14/17 (82.4%) | Microbiological identification and AST performed using automated Vitek-2 system and E-test. Results interpreted according to CLSI criteria. |
| Nasim et al. (2023) | Retrospective, single centre, case-control study | Pakistan | 1677 Renal transplant recipients between 2015 -2019 | 3GCR-E  CRPA | 3GCR-E 27/36 (75%)  CRPA 1/4 (25%) |  | Microbiological identification and AST methods ot described. Susceptibility results interpreted according to CLSI criteria. |
| Neofytos et al. (2023) | Retrospective, nested, multicentre, cohort study | Switzerland | 4383 SOT recipients (359 heart, 2437 kidney, 1044 liver, 441 lung, 102 kidney-pancreas) from 2008 to 2019, using the Swiss Transplant Cohort Study registry | MRSA  VRE  3GCR-E  CRPA | 557 episodes of BSI among 415 SOT recipients  MRSA 7/28 (25%)  VRE 2/67 (3.0%)  3GCR-E 32/301 (10.6%)  CRPA 0/59 (0%) | Not reported | Not reported |
| Nie et al. (2015) | Retrospective, multicentre, observational analysis (2 sites) | China | 1850 SOT recipients (1557 kidney, 282 liver, 3 heart, 7 liver-kidney, 1 kidney-pancreas) between January 2003 - April 2015 | CRAB | CRAB 21/36 (58.3%) | Not reported | Microbiological identification was performed using the Vitek-2 automated system. AST was conducted using the Kirby-Bauer disc diffusion method and MIC tests according to CLSI criteria. |
| Oriol et al. (2015) | Prospective, single centre, observational study | Spain | 361 consecutive episodes of BSI among 246 SOT recipients (kidney, liver, heart and multivisceral) between January 2007 - October 2014 | MRSA  VRE  3GCR-E | 361 episodes of BSIs  MRSA 4/19 (21.1%)  VRE 0/11 (0.0%)  3GCR-E 34/175 (19.4%) | Not reported | Microbial identification and AST were performed using either Microscan or Vitek automated systems. Results were interpreted according to CLSI criteria. |
| Oriol et al. (2017) | Prospective, single centre, cohort study | Spain | 1829 adult SOT recipients (1113 kidney, 547 liver, 169 heart) between January 2007 - December 2016 | 3GCR-E | 475 episodes of BSI; 218 in the first year after transplant.  3GCR-E 44/105 (41.9%) | Not reported | Microbiological identification performed using either Microscan or Vitek automated systems. Results were interpreted according to EUCAST criteria. |
| Sganga et al. (2012) | Retrospective, single centre, cohort study | Italy | 75 liver transplant patients between January 2008 - July 2011 | MRSA  VRSA  3GCR-E | 21 episodes of BSI  MRSA 1/1 (100%)  VRSA 0/1 (0%)  3GCR-E 4/15 (26.7%) | Not reported | Microbiological confirmation and AST were performed using the Vitek-2 automated system. The presence of genes encoding -lactamases was evaluated by PCR amplification and sequencing. Results were interpreted according to CLSI criteria. |
| Shendi et al. (2016) | Retrospective, single centre, observational study | United Kingdom | 1152 kidney transplant recipients between July 2009 - April 2016 | 3GCR-E | 116 BSI episodes among 87 patients  3GCR-E – 14/75 (18.7%) | Not reported | Microbiological identification and AST were performed using the BD Phoenix automated system until October 2011 then transitioned to the Bruke MALDI-ToF system. Results were interpreted according to EUCAST criteria. |
| Shi et al. (2009) | Prospective, single centre, case control study | China | 475 liver transplant recipients between January 2003 - December 2006 | 3GCR-E  CRE  CRAB | 255 episodes of Gram negative BSI among 152 patients  3GCR-E 25/42 (59.5%)  CRE 4/42 (9.5%)  CRAB 18/30 (60%) | Not reported | Microbiological species identification was conducted using the Vitek automated system. AST was performed using the Kirby-Bauer disc diffusion method and interpreted according to NCCLS (CLSI) criteria. |
| Shi et al. (2010) | Prospective, single centre, cohort study | China | 475 liver transplant recipients between January 2003 - December 2006 | MRSA/ VISA  VRE | 98 episodes of BSI among 82 patients.  MRSA 13/13 (100%)  VRSA - 0/13 (0%)  VRE 0/34 (0%) | Not reported | Microbiological identification was conducted using the Vitek automated system. AST was performed using the Kirby-Bauer disc diffusion method and interpreted using NCCLS (CLSI) criteria. |
| Simkins et al. (2014) | Retrospective, single centre, case control study | United States | 522 kidney transplant recipients who underwent transplantation between 1  January 2006 and 31 December 2010  Excluded multivisceral (e.g. kidney-liver transplants) | CRE (K pneumoniae only) | CRE (K. pneumoniae) - 5/11 (45.5%) | Not reported | Not reported |
| Simkins et al. (2019) | Retrospective, single centre, observational study | United States | 55 intestinal transplant patients who developed BSI due to enteric organisms within the first 6 months of transplannt, between January 2009 and May 2017 | VRE  3GCR-E  CRE | Total 51 episodes of BSIs among 28 patients  VRE 14/14 (100%)  3GCR-E 6/22 (27.3%)  CRE 3/22 (14.2%) | Not reported | Not reported |
| Singh et al. (2004) | Retrospective, single centre, observational study | United States | 233 liver transplant recipients between 1989 – 2003 | MRSA  VRE | Not reported | MRSA mortality 9/33 (27.3%)  VRE mortality 4/8 (50%) | Not reported |
| Spence et al. (2021) | Retrospective, single centre, observational study | United States | 106 intestinal and multivisceral transplants among 103 recipients between 2003 and 2015 | VRE  CRE (but no denominator) | 62 transplant recipients who developed a BSI    VRE 17/36 (47.2%)  CRE – 0% | Not reported | Not reported |
| Torre-Cisneros et al. (2002) | Retrospective, multicentre (3 sites) observational study | Spain | 392 Liver transplant recipients between Jan 1994 - October 1999 | MRSA | MRSA 5/14 (35.7%) | Not reported | Not reported |
| Tran-Dinh et al. (2022) | Retrospective, single centre, observational study | France | 303 lung transplant recipients with ICU admission between January 2015 - October 2021 | MRSA  3GCR-E | 45 episodes of BSI among 33 patients  MRSA = 0/8 (0%)  3GCR-E = 6/11 (54.5%) | Not reported | Microbiological identification was performed using the MALDI-ToF system and AST performed using double disc diffusion method. Results were interpreted according to EUCAST criteria. |
| Wu et al. (2020) | Retrospective, single centre cohort study | China | 1249 ASOT recipients (1039 kidney, 210 liver) between December 2012 -July 2019 | CRE (K. pneumoniae only) | CRE 16/24 | Not reported | Microbiological species identification was conducted using the Vitek automated system. AST was performed using the Kirby-Bauer disc diffusion method and interpreted according to CLSI criteria. |
| Wu et al. (2020) | Retrospective, single centre observational study. | China | 1179 abdominal SOT recipients (977 kidney, 202 liver) from October 2013 - June 2019. | CRE (K. pneumoniae only) | CRE 13/21 | Not reported | Microbiological species identification was conducted using the Vitek automated system. AST was performed using the Kirby-Bauer disc diffusion method and interpreted according to CLSI criteria. |
| Ye et al. (2014) | Retrospective, multicentre observational study (2 sites) | China | 71 SOT recipients (liver, kidney, heart, multivisceral) between January 2002 - August 2013 | MRSA  VRE  3GCR-E  CRAB  CRPA | 84 episodes of BSI among 71 SOT recipients.   VRE 0/13 (0%)  MRSA 18/26 (69.2%)  3GCR-E 10/15 (33.3%)  CRAB 10/20 (50%)  CRPA 3/10 (30%) | Pooled MDRO 30-day mortality 22/39 (56.4%)  Pool non-MDRO 30-day mortality 12/32 (37.5%) | Microbiological species identification was conducted using the Vitek automated system. AST was performed using the Kirby-Bauer disc diffusion method and interpreted according to NCCLS criteria. |
| Zhou et al. | Retrospective, multicentre, observational study (2 sites) | China | 275 liver transplant recipients with S. aureus BSI between January 2001 -December 2014 | MRSA | MRSA 16/20 (80%) | MRSA mortality 9/20 (45%) | Microbiological species identification was conducted using the Vitek automated system. AST was performed using the Kirby-Bauer disc diffusion method and interpreted according to NCCLS criteria. |

**Abbreviations:**

3GCR-E: Third-generation cephalosporin-resistant Enterobacterales

AST: Antimicrobial susceptibility testing

BSI: Bloodstream infection

CLSI: Clinical & Laboratory Standards Institute

CRAB: Carbapenem-resistant *Acinetobacter baumannii*

CRE: Carbapenem-resistant Enterobacterales

CRPA: Carbapenem-resistant *Pseudomonas aeruginosa*

EUCAST: European Committee on Antimicrobial Susceptibility Testing

MRSA: Methicillin-resistant *Staphylococcus aureus*

NCCLS: National Committee for Clinical Laboratory Standards

SOT: Solid organ transplant

VRE: Vancomycin-resistant *Enterococcus faecium*

**Supp. Table 4 – Newcastle-Ottawa Scale (NOS) Quality Assessment Form**

| Selection | 1) Representativeness of the exposed cohort **– Solid organ transplant recipient with multidrug resistant bloodstream infection** | a) Truly representative (one star) b) Somewhat representative (one star) c) Selected group d) No description of the derivation of the cohort |
| --- | --- | --- |
|  | 2) Selection of the non-exposed cohort – **Solid organ transplant recipient with drug-susceptible bloodstream infection** | a) Drawn from the same community as the exposed cohort (one star) b) Drawn from a different source c) No description of the derivation of the non-exposed cohort |
|  | 3) Ascertainment of exposure | a) Secure record (e.g., surgical record) (one star) – **microbiological confirmation of positive blood culture** b) Structured interview (one star) c) Written self report d) No description e) Other |
|  | 4) Demonstration that outcome of interest was not present at start of study – **baseline blood cultures** | a) Yes (one star) b) No |
| Comparability | 1) Comparability of cohorts on the basis of the design or analysis controlled for confounders | a) The study controls for age, sex and **baseline demographics** (one star) b) Study controls for other factors (list) - **immunosuppression regimen** (one star) c) Cohorts are not comparable on the basis of the design or analysis controlled for confounders |
| Outcome | 1) Assessment of outcome | a) Independent blind assessment (one star) – **microbiological confirmation with species identification and antimicrobial susceptibility methods stated** b) Record linkage (one star) c) Self report d) No description e) Other |
|  | 2) Was follow-up long enough for outcomes to occur | a) Yes (one star) b) No |
|  | 3) Adequacy of follow-up of cohorts   Indicate the median duration of follow-up and a brief rationale for the assessment above: **All follow-up periods included** | a) Complete follow up- all subject accounted for (one star) b) Subjects lost to follow up unlikely to introduce bias- number lost less than or equal to 20% or description of those lost suggested no different from those followed. (one star) c) Follow up rate less than 80% and no description of those lost d) No statement |

E-18

Thresholds for converting the Newcastle-Ottawa scales to AHRQ standards (good, fair, and poor):

Good quality: 3 or 4 stars in selection domain AND 1 or 2 stars in comparability domain AND 2 or 3 stars in outcome/exposure domain

Fair quality: 2 stars in selection domain AND 1 or 2 stars in comparability domain AND 2 or 3 stars in outcome/exposure domain

Poor quality: 0 or 1 star in selection domain OR 0 stars in comparability domain OR 0 or 1 stars in outcome/exposure domain

**Supp. Table 5 – Newcastle Ottawa Scale (NOS) scores.**

| **Study** | **Selection** | | | | **Comparability** | | **Outcome** | | | **Total (max. score 9)** |
| --- | --- | --- | --- | --- | --- | --- | --- | --- | --- | --- |
|  | Representative of the exposed cohort | Selection of non-exposed cohort | Ascertainment of exposure | Outcome of interest not present at start of the study | Baseline demographics | AST methods | Assessment of outcomes | Sufficient follow up time | Adequacy of follow up |  |
| Adelman et al. (2024) | 1 | 1 | 1 | 0 | 1 | 1 | 1 | 1 | 1 | 8 |
| Aguiar et al. (2014) | 1 | 1 | 1 | 0 | 1 | 1 | 1 | 1 | 1 | 8 |
| Anesi et al. (2021) | 1 | 1 | 1 | 0 | 1 | 1 | 1 | 1 | 1 | 8 |
| Anesi et al. (2023) | 1 | 1 | 1 | 0 | 1 | 1 | 1 | 1 | 1 | 8 |
| Basaran et al. (2022) | 1 | 1 | 1 | 0 | 1 | 1 | 1 | 1 | 1 | 8 |
| Bedini et al. (2007) | 1 | 1 | 1 | 0 | 1 | 0 | 1 | 1 | 1 | 7 |
| Berger et al. (2006) | 1 | 1 | 1 | 0 | 1 | 0 | 1 | 1 | 1 | 7 |
| Bert et al. (2010) | 1 | 1 | 1 | 0 | 1 | 1 | 1 | 1 | 1 | 8 |
| Bodro et al. (2013) | 1 | 1 | 1 | 0 | 1 | 1 | 1 | 1 | 1 | 8 |
| Camargo et al. (2015) | 1 | 1 | 1 | 0 | 1 | 1 | 1 | 1 | 1 | 8 |
| Chueiri Neto et al. (2019) | 1 | 1 | 1 | 0 | 1 | 0 | 1 | 1 | 1 | 7 |
| Dasakalaki et al. (2014) | 1 | 1 | 1 | 0 | 1 | 1 | 1 | 1 | 1 | 8 |
| De Gouvea et al. (2012) | 1 | 1 | 1 | 0 | 1 | 1 | 1 | 1 | 1 | 8 |
| Dubler et al. (2020) | 1 | 1 | 1 | 0 | 1 | 1 | 1 | 1 | 1 | 8 |
| Eichenberger et al. (2021) | 1 | 1 | 1 | 0 | 1 | 1 | 1 | 1 | 1 | 8 |
| Gagliotti et al. (2018) | 1 | 1 | 1 | 0 | 1 | 0 | 1 | 1 | 1 | 7 |
| Gao et al. (2015) | 1 | 1 | 1 | 0 | 1 | 1 | 1 | 1 | 1 | 8 |
| Hashimoto et al. (2008) | 1 | 1 | 1 | 0 | 1 | 1 | 1 | 1 | 1 | 8 |
| Husain et al. (2006) | 1 | 1 | 1 | 0 | 1 | 0 | 1 | 1 | 1 | 7 |
| Iida et al. (2010) | 1 | 1 | 1 | 0 | 1 | 0 | 1 | 1 | 1 | 7 |
| Karruli et al. (2021) | 1 | 1 | 1 | 0 | 1 | 1 | 1 | 1 | 1 | 8 |
| Kim et al. (2009) | 1 | 1 | 1 | 0 | 1 | 1 | 1 | 1 | 1 | 8 |
| Kim et al. (2013) | 1 | 1 | 1 | 0 | 1 | 1 | 1 | 1 | 1 | 8 |
| Kim et al. (2018) | 1 | 1 | 1 | 0 | 1 | 1 | 1 | 1 | 1 | 8 |
| Kim et al. (2019) | 1 | 1 | 1 | 0 | 1 | 1 | 1 | 1 | 1 | 8 |
| Kim et al. (2021) | 1 | 1 | 1 | 0 | 1 | 1 | 1 | 1 | 1 | 8 |
| Lee et al. (2011) | 1 | 1 | 1 | 0 | 1 | 1 | 1 | 1 | 1 | 8 |
| Lee et al. (2022) | 1 | 1 | 1 | 0 | 1 | 1 | 1 | 1 | 1 | 8 |
| Luo et al. (2016) | 1 | 1 | 1 | 0 | 1 | 1 | 1 | 1 | 1 | 8 |
| Malinis et al. (2012) | 1 | 1 | 1 | 0 | 1 | 1 | 1 | 1 | 1 | 8 |
| Mercuro et al. (2020) | 1 | 1 | 1 | 0 | 1 | 1 | 1 | 1 | 1 | 8 |
| Min et al. (2023) | 1 | 1 | 1 | 0 | 1 | 0 | 1 | 1 | 1 | 7 |
| Mouloudi et al. (2014) | 1 | 1 | 1 | 0 | 1 | 1 | 1 | 1 | 1 | 8 |
| Nasim et al. (2023) | 1 | 1 | 1 | 0 | 1 | 1 | 1 | 1 | 1 | 8 |
| Neofytos et al. (2023) | 1 | 1 | 1 | 0 | 1 | 0 | 1 | 1 | 1 | 7 |
| Nie et al. (2015) | 1 | 1 | 1 | 0 | 1 | 1 | 1 | 1 | 1 | 8 |
| Oriol et al. (2015) | 1 | 1 | 1 | 0 | 1 | 1 | 1 | 1 | 1 | 8 |
| Oriol et al. (2017) | 1 | 1 | 1 | 0 | 1 | 1 | 1 | 1 | 1 | 8 |
| Sganga et al. (2012) | 1 | 1 | 1 | 0 | 1 | 1 | 1 | 1 | 1 | 8 |
| Shendi et al. (2016) | 1 | 1 | 1 | 0 | 1 | 1 | 1 | 1 | 1 | 8 |
| Shi et al. (2009) | 1 | 1 | 1 | 1 | 1 | 1 | 1 | 1 | 1 | 9 |
| Shi et al. (2010) | 1 | 1 | 1 | 0 | 1 | 1 | 1 | 1 | 1 | 8 |
| Simkins et al. (2014) | 1 | 1 | 1 | 0 | 1 | 0 | 1 | 1 | 1 | 7 |
| Simkins et al. (2019) | 1 | 1 | 1 | 0 | 1 | 0 | 1 | 1 | 1 | 7 |
| Singh et al. (2004) | 1 | 1 | 1 | 0 | 1 | 1 | 1 | 1 | 1 | 8 |
| Spence et al. (2021) | 1 | 1 | 1 | 0 | 1 | 0 | 1 | 1 | 1 | 7 |
| Torre-Cisneros et al. (2002) | 1 | 1 | 1 | 0 | 1 | 0 | 1 | 1 | 1 | 7 |
| Tran-Dinh et al. (2022) | 0 | 1 | 1 | 0 | 1 | 1 | 1 | 1 | 1 | 7 |
| Wu et al. (2020) | 1 | 1 | 1 | 0 | 1 | 1 | 1 | 1 | 1 | 8 |
| Wu et al. (2020) | 1 | 1 | 1 | 0 | 1 | 1 | 1 | 1 | 1 | 8 |
| Ye et al. (2014) | 1 | 1 | 1 | 0 | 1 | 1 | 1 | 1 | 1 | 8 |
| Zhou et al. (2015) | 1 | 1 | 1 | 0 | 1 | 1 | 1 | 1 | 1 | 8 |

**Supp. Table 6 – Linear regression analysis results for specific multi-drug resistant organism trends over time.**

| **Multi-drug resistant organism** | **Intercept** | **Slope** | **Standard error** | **t value** | ***P* value** | **R-squared** |
| --- | --- | --- | --- | --- | --- | --- |
| 3GCR-E | 2.832401 | -0.001207 | 0.009103 | -0.133 | 0.896 | 0.0009757 |
| CRAB | -47.48543 | 0.02388 | 0.01821 | 1.311 | 0.219 | 0.1468 |
| CRE | 9.704469 | -0.004687 | 0.018450 | -0.254 | 0.805 | 0.007121 |
| CRPA | 54.44768 | -0.02681 | 0.01631 | -1.644 | 0.135 | 0.2308 |
| MRSA | 32.659667 | -0.015928 | 0.009755 | -1.633 | 0.116 | 0.1039 |
| VRE | 4.444150 | -0.001997 | 0.016066 | -0.124 | 0.902 | 0.0008123 |

**Abbreviations:**

3GCR-E: Third-generation cephalosporin-resistant Enterobacterales

CRAB: Carbapenem-resistant *Acinetobacter baumannii*

CRE: Carbapenem-resistant Enterobacterales

CRPA: Carbapenem-resistant *Pseudomonas aeruginosa*

MRSA: Methicillin-resistant *Staphylococcus aureus*

VRE: Vancomycin-resistant *Enterococcus faecium*
